# Tracking private WhatsApp discourse about COVID-19: A longitudinal infodemiology study in Singapore

**DOI:** 10.1101/2020.09.29.20203646

**Authors:** Edina YQ Tan, Russell RE Wee, Saw Young Ern, Kylie JQ Heng, Joseph WE Chin, Eddie M.W. Tong, Jean CJ Liu

## Abstract

**Background:** Worldwide, social media traffic increased following the onset of the coronavirus disease (COVID-19) pandemic. Although the spread of COVID-19 content has been described for several social media platforms (e.g., Twitter, Facebook), little is known about how content is spread via private messaging platforms such as WhatsApp.

**Objective:** In this study, we documented: (i) how WhatsApp is used to transmit COVID-19 content; (ii) the characteristics of WhatsApp users based on their usage patterns; and (iii) how usage patterns link to well-being.

**Methods:** We used the experience sampling method to track day-to-day WhatsApp usage during the COVID-19 pandemic. For one week, participants reported each day the extent to which they had received, forwarded, or discussed COVID-19 content. The final dataset comprised 924 data points collected from 151 participants.

**Results:** During the week-long monitoring, most participants (143/151, 95%) reported at least one COVID-19-related use of WhatsApp. When a taxonomy was generated based on usage patterns, 1 in 10 participants (21/151, 14%) were found to have received and shared a high volume of forwarded COVID-19 content – akin to ‘super spreaders’ identified on other social media platforms. Finally, those who engaged with more COVID-19 content in their personal chats were more likely to report having COVID-19 thoughts throughout the day.

**Conclusions:** These findings provide a rare window into discourse on private messenger platforms. In turn, this can inform risk communication strategies during the pandemic.

## Introduction

WhatsApp is the most commonly used messaging application worldwide, with 1.5 billion users across 180 countries [1]. On account of its large user base and near-instant message transmission, the platform has played a critical role in risk communication during the coronavirus disease (COVID-19) pandemic.

On the one hand, WhatsApp has been co-opted by government agencies and the World Health Organization to disseminate official COVID-19 updates [2]. While this showcases the platform’s ability to reach a large sector of the population, the same feature has also made it a vessel for misinformation. For example, at the beginning of the pandemic, WhatsApp noted a 40% surge in usage [3]. This was paired with a high volume of message-forwarding activity widely believed to support misinformation, leading the platform to restrict the number of individuals a message could be forwarded to simultaneously [4,5].

Despite these restrictions, a survey in India found that 1 in 2 participants had received COVID-19 misinformation through WhatsApp or Facebook [6]. Likewise, WhatsApp was identified by Hong Kong residents as the foremost source for COVID-19 rumors [7]. As misinformation can jeopardize public health strategies, these findings underscore the need for infodemiological studies documenting how COVID-19 content spreads through WhatsApp.

To date, however, the bulk of infodemiology studies have focused on social media platforms where content is publicly accessible (e.g., Twitter, Facebook) [8,9]. By contrast, research on WhatsApp has proven elusive because of its private nature, with end-to-end encryption ensuring that only senders and recipients have access to messages sent through the platform. Nonetheless, WhatsApp research remains a priority: aside from its popularity and role in carrying crisis-related misinformation [10,11], insights from public posts are also unlikely to generalize to WhatsApp’s private messages [12]. It thus remains unclear who sends COVID-19 messages, to whom, and in what manner.

Addressing these gaps in the literature, we designed a study to: (i) describe the base rate of COVID-19 content dissemination; (ii) understand WhatsApp users; and (iii) examine correlates of usage patterns. Specifically, we applied the experience sampling method to track one week of WhatsApp usage amidst everyday routines [13,14], asking participants to report each day their frequency of receiving, forwarding, or discussing COVID-19-related content. Through this method, we generated a taxonomy of participants based on their usage patterns, and examined whether day-to-day variations in WhatsApp usage predicted COVID-19 concerns.

## Methods

### Recruitment

Between 17 March to 7 May 2020, participants were recruited from the general community via advertisements placed in Facebook and WhatsApp community groups (e.g., residential groups, workplace groups, university groups), posts on popular online forums, and paid Facebook advertisements targeting Singapore-based users. All study activities took place via the online survey platform Qualtrics, and participants were reimbursed SGD $5 upon study completion. The study protocol was approved by the Yale-NUS College Ethics Review Committee (2020-CERC-001) and was pre-registered at ClinicalTrials.gov (NCT04367363).

### Participants

Participants were 151 adults who met the following inclusion criteria: (1) aged 21 years or older, (2) had lived in Singapore for at least 2 years, and (3) had a WhatsApp account.

### Measures

Following informed consent, participants completed: (i) a baseline questionnaire; (ii) experience sampling responses daily for 7 days; and (iii) a final questionnaire (Figure 1).

**Figure 1.**
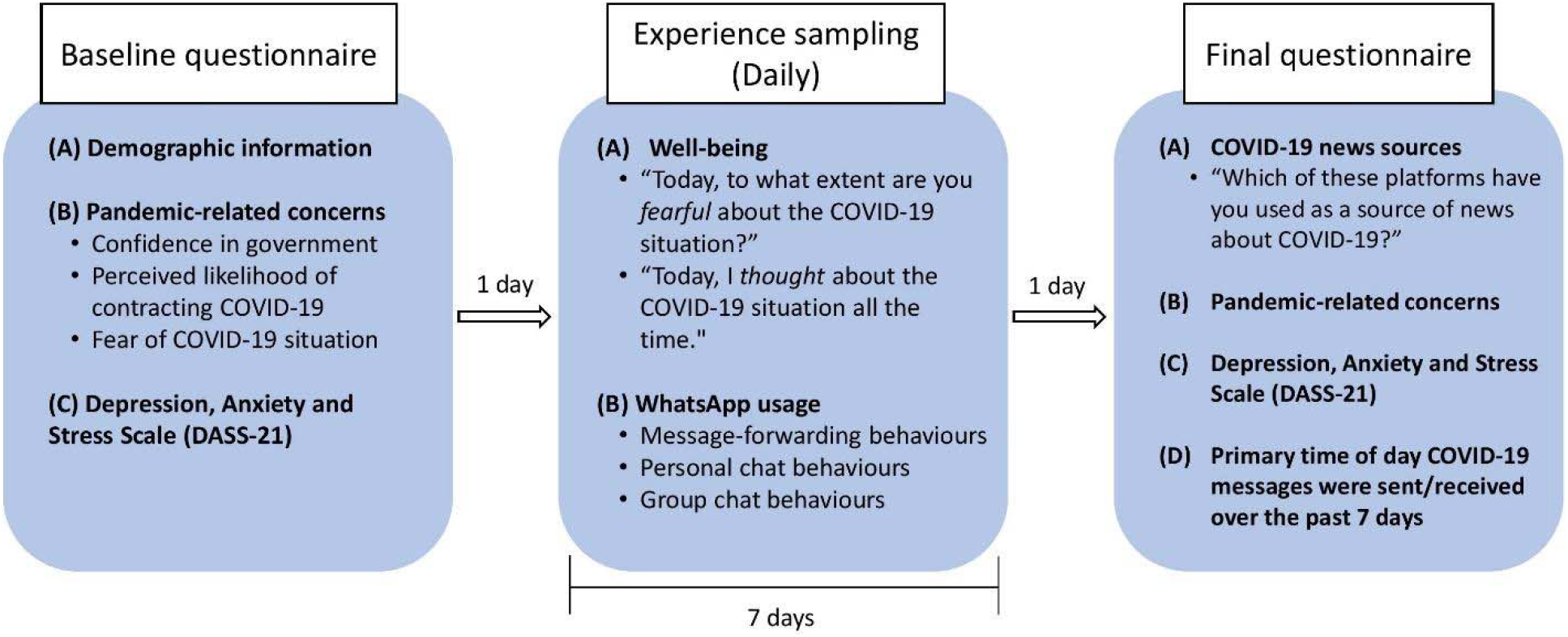
Schematic of study procedures. All participants completed a baseline questionnaire, followed by 7 days of experience sampling where participants addressed questions about well-being and WhatsApp usage daily. Participants completed a final questionnaire one day after the experience sampling protocol ended.

#### Experience sampling

As the primary form of data collection, we used the experience sampling method to capture “COVID-19 chatter” on WhatsApp. Through this protocol, we collected 924 data points across 151 participants (compliance rate: 86.2%).

For 7 days, participants accessed an online survey each evening (2130 hrs) to report WhatsApp usage for the day. First, participants indicated whether they had forwarded messages related to COVID-19 (yes/no). We focused on message-forwarding as a proxy indicator for high-risk content, since: (i) large Twitter studies have observed that misinformation is more likely to be shared than posts that are true [15], and (ii) because WhatsApp developers had previously linked forwarded messages to misinformation [4,5]. If participants had forwarded COVID-19 content, they were then asked: (i) how many unique COVID-19 messages they had forwarded, and (ii) how many unique groups and individuals they had forwarded messages to.

Additionally, participants were asked about their personal chats – their one-to-one chats on WhatsApp. They indicated whether COVID-19 messages had been forwarded to them in personal chats (yes/no), and, if so: (i) how many unique messages they had received, and (ii) how many different people they had received messages from. Thereafter, participants recounted whether they had discussed COVID-19 in conversations where either they or the other party generated message(s) related to COVID-19 (yes/no), and – if so – how many unique chats were involved.

Finally, for group chats, participants were asked if COVID-19 had been mentioned in any of their WhatsApp groups by at least one other person (not including themselves) (yes/no). This could have occurred either through others forwarding messages, or through others generating their own comments. Affirmative responses were followed with a question on how many WhatsApp groups had done so.

Aside from WhatsApp metrics, participants also reported their COVID-19 concerns for the day: (i) how afraid they felt about the COVID-19 situation (4-point scale with 1= “Not scared at all” and 4=“Very scared”); and (ii) whether they had thought about the COVID-19 situation all the time (5-point scale with 1=“Not at all true” and 5=“Very true”).

#### Baseline and final questionnaires

To characterize participants, we included baseline and final questionnaires where participants reported: demographics (age, gender, religion, ethnicity, marital status, education, house type, household size, citizenship, country of birth, and years in Singapore), the time of day they read and sent COVID-19 messages on WhatsApp (mostly in the morning, afternoon, evening, late night, or throughout the day), and sources through which they obtained COVID-19 news (e.g., printed newspapers, radio, WhatsApp, YouTube). Additionally, participants reported their well-being (using the 21-item Depression, Anxiety and Stress Scale (DASS-21; [16]), and responses to the pandemic [2,17]: (1) how confident they were that the government could control the nationwide spread of COVID-19 (1 = “Not confident at all, 4 = “Very confident”); (2) how likely they judged that they (or someone in their immediate household) would be infected with COVID-19 (1 = “Not at all likely”, 4 = “Very likely”); and (3) how fearful they were about the situation in the country (1 = “Not scared at all”, 4 = “Very scared”).

### Statistical Analysis

We first summarized the data with counts (%) or means (standard deviations), focusing on the 7 quantitative WhatsApp usage variables (Figure 2): the number of (1) COVID-19 messages participants forwarded; (2) groups forwarded to; (3) individuals forwarded to; (4) forwarded messages received; (5) individuals from whom messages were received; (6) personal chats involving COVID-19 conversations; and (7) group chats discussing COVID-19. (Multimedia Appendix 1 shows the pattern of correlations across these variables).

**Figure 2.**
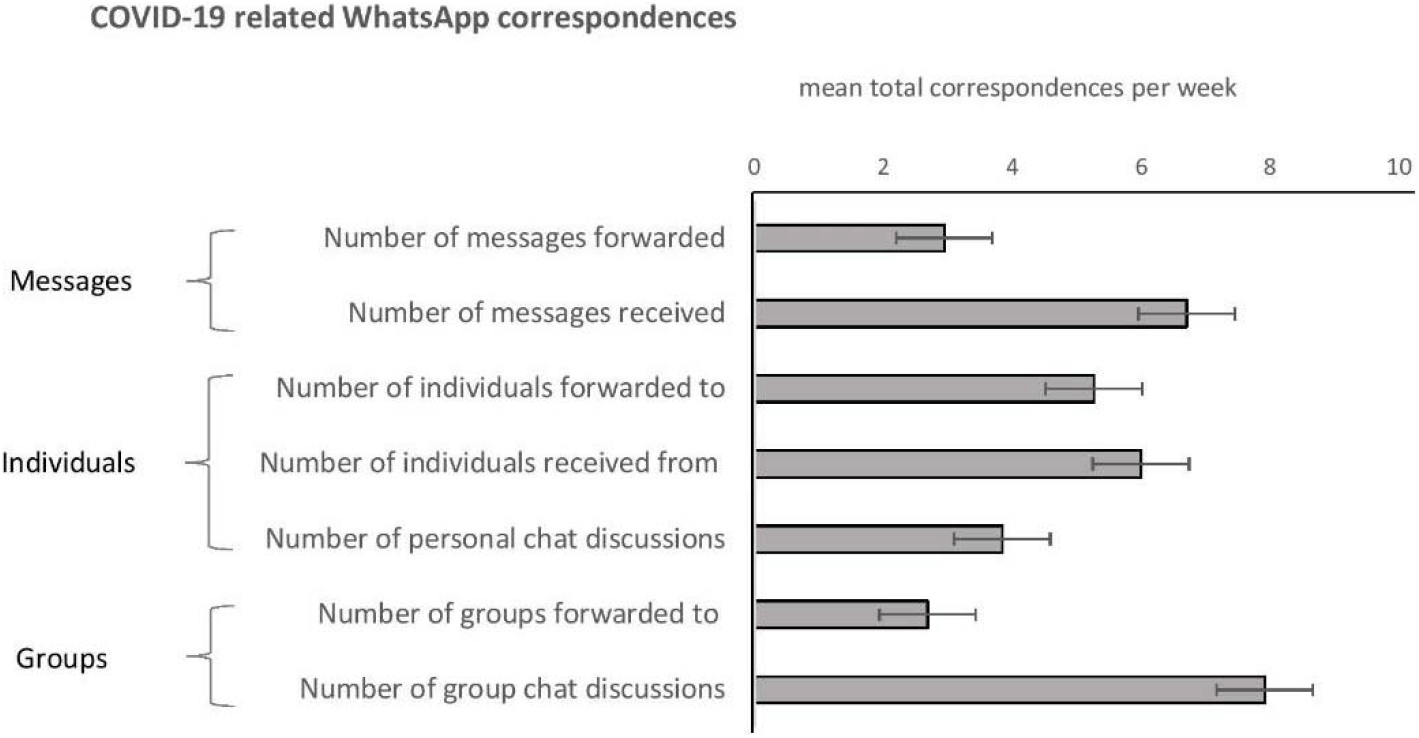
Distribution of COVID-19 related WhatsApp behaviors. In a week-long experience sampling procedure, participants reported the extent to which they engaged in COVID-19 WhatsApp behaviors (either by forwarding or receiving messages, or in conversations). Horizontal bars represent the total amount of each activity captured, averaged across all participants. Horizontal lines represent the 95% confidence interval for the mean.

Next, to understand user profiles, we applied latent profile analysis to create a taxonomy of participants based on their WhatsApp usage (R package: ‘mclust’ [18]). Latent profile analysis is a bottom-up statistical clustering method that defines classes of people based on common characteristics. Using all observations of a continuous dependent variable, classes are created such that within each class, indicator variables are statistically uncorrelated [19]. We thus applied this technique to cluster participants based on their responses to the 7 WhatsApp usage variables, with values obtained by aggregating the reported frequency of each variable over the week. To uncover clusters, we used Gaussian Mixture Models (GMM) and assigned cluster membership using Bayesian probabilities. The final number of clusters was determined using the Bayesian Information Criterion (BIC), Integrated Completed Likelihood Criterion (ICL), and a Bootstrap Likelihood Test (BLRT).

Finally, we examined whether day-to-day variations in COVID-19 WhatsApp chatter was tracked by variations in COVID-19 concerns. As predictors, we first quantified COVID-19 chatter on personal and group chats. For personal chats, the following variables were summed within each day and for each participant: the number of (i) individuals COVID-19 messages were forwarded to; (ii) individuals from whom forwarded messages were received; and (iii) personal conversations discussing COVID-19. For group chats, the following variables were summed: the number of (i) groups participants forwarded COVID-19 messages to; and (ii) groups where COVID-19 messages were mentioned. Scores were grand-mean centered by subtracting the mean number of chats across subjects and time points from each score (*M =* 2.47 and *M* = 1.29 respectively). In addition, we created between- and within-subject versions of each predictor [20]. The final analyses involved linear mixed-effects models for each outcome measure (fear and thoughts about COVID-19), with the following entered as fixed effects: time (centered such that 0 referred to the middle of the week), daily personal chats (between-subjects), daily personal chats (within-subjects), daily group chats (between-subjects), and daily group chats (within-subjects). Random intercepts accounted for correlated data due to repeated measures.

Across all analyses, the type 1 decision-wise error rate was controlled at α = 0.05. All statistical analyses were conducted in R 3.5.0 [21] and SPSS 25 [22].

## Results

### Baseline Participant Characteristics

As shown in Table 1, 69% (104/151) of participants identified as female, with a mean age of 36.35 years (SD 14.7). Participants were predominantly of Asian ethnicity (93% Chinese; 140/151), and had at least post-secondary education (88.1%, 133/151). 40% (60/151) were married, and the majority (69.5%, 105/151) belonged to households of at least 4 members.

**Table 1.**
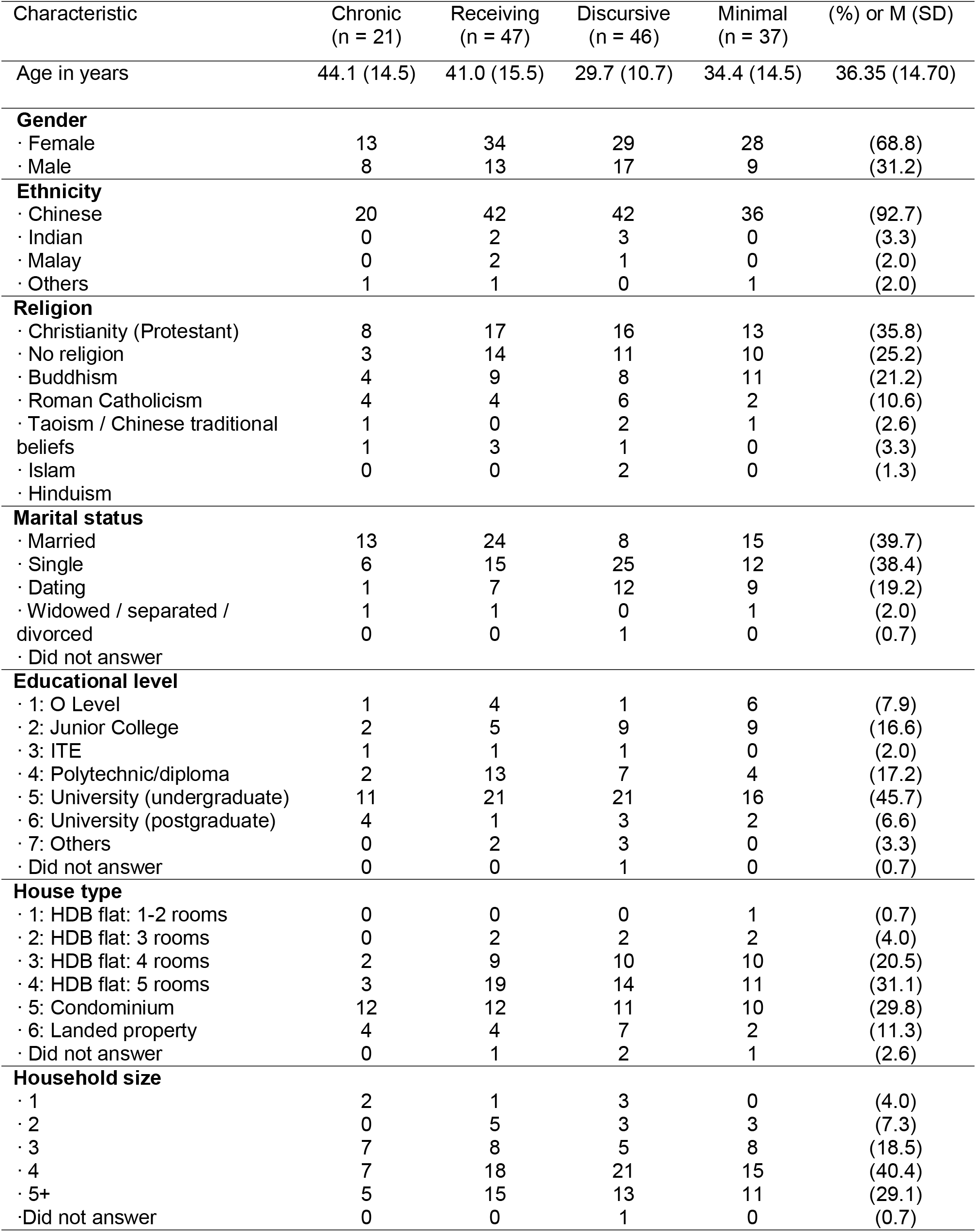
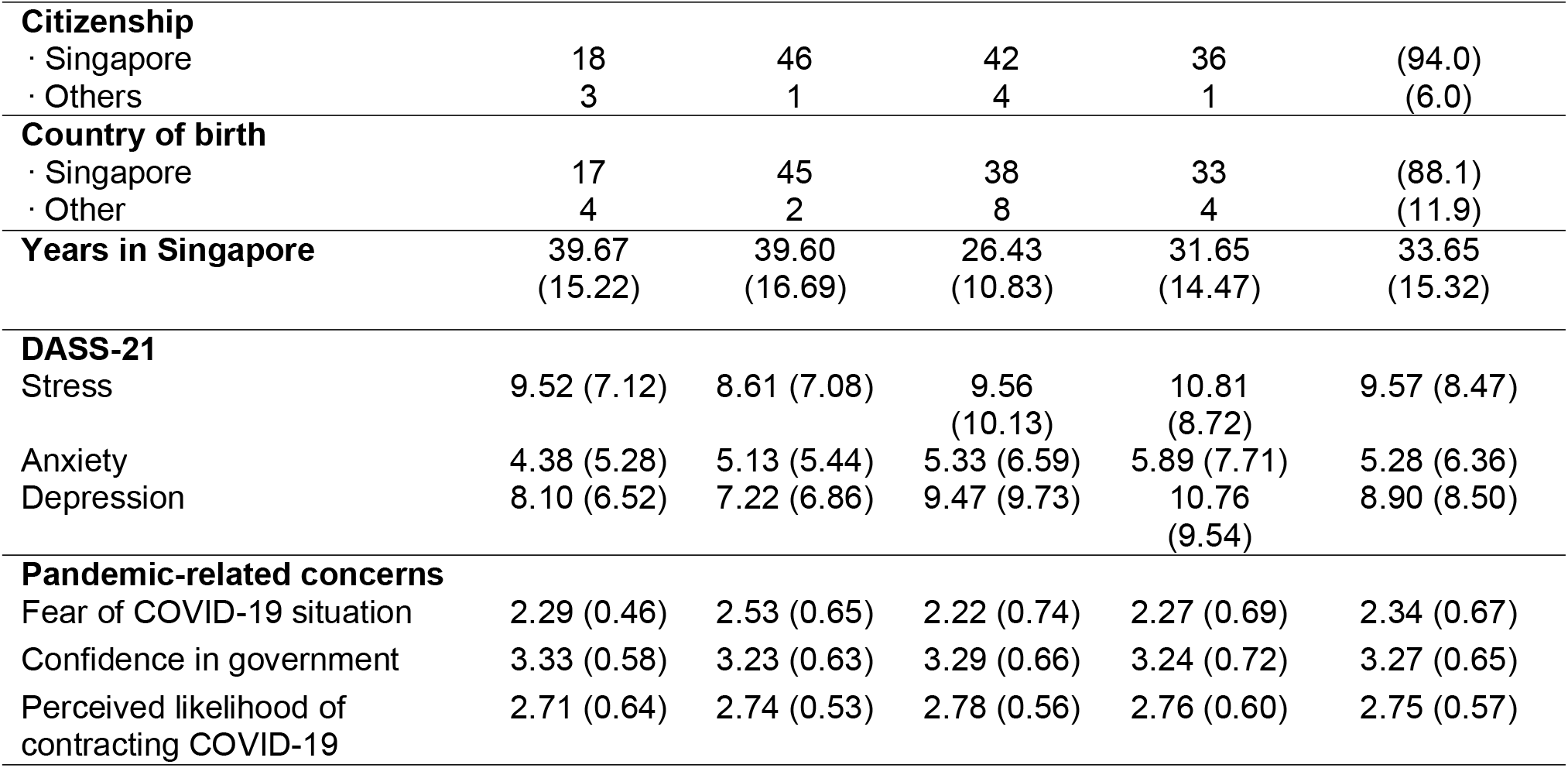
Participant characteristics as a function of COVID-19 WhatsApp usage patterns.

### Base rate of COVID-19 WhatsApp usage

Participants’ self-reports revealed that WhatsApp was the second-most common source for COVID-19 news, after news website or apps (Figure 3). Quantifying this through one week of experience sampling, we found that nearly all participants (95%, 143/151; 95% CI: 90-98%) reported at least one COVID-19 related use of WhatsApp. Namely, 1 in 2 participants (52%, 79/151; 95% CI: 44-60%) forwarded at least one COVID-19 message (to either individuals or groups), 78% (118/151; 95% CI: 71 - 84%) received at least one forwarded message in personal chats, 66% (100/151; 95% CI: 58-74%) engaged in personal chat conversations about COVID-19, and 88% (133/151; 95% CI: 82% - 93%) had been in groups where COVID-19 was mentioned.

**Figure 3.**
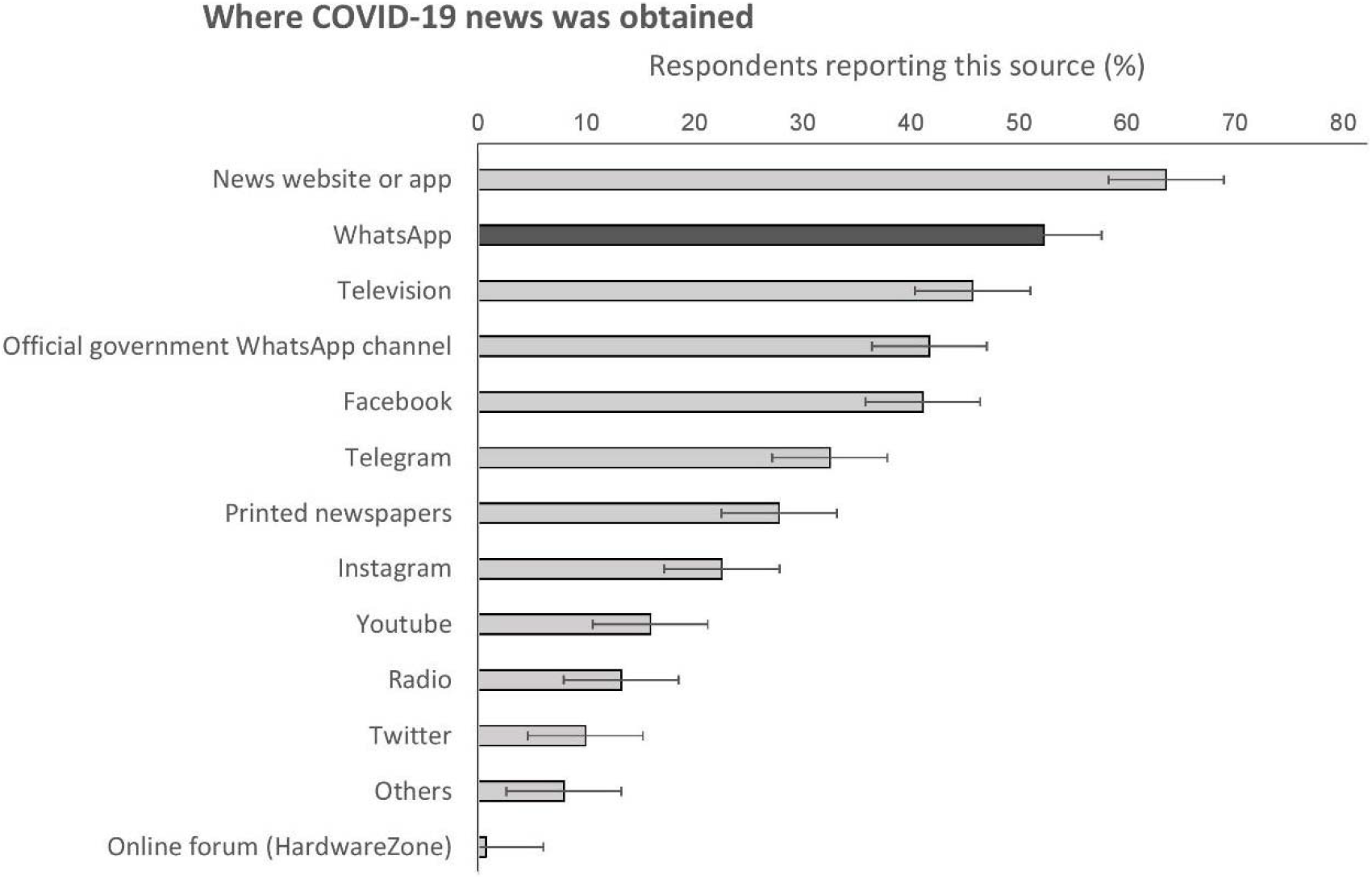
Sources of COVID-19 news. In a questionnaire, participants self-reported where they received COVID-19 news from.

Figure 2 shows the extent to which participants engaged in each of these activities. On average, participants: (i) received two times more messages (2.3) than they forwarded on, and (ii) were more likely to forward messages to individuals than to groups (average of 5.3 versus 2.7 messages a week, respectively). Beyond passive engagement, participants also took part in an average of 3.8 one-to-one conversations about COVID-19 during the week; however, these interactions occurred less frequently than the sending or receiving of forwarded messages in group chats.

### Characterizing participants based on COVID-19 WhatsApp usage

#### Latent profile analysis: Generating a taxonomy of WhatsApp usage

Although most participants received and shared COVID-19 content on WhatsApp, there were individual differences in usage patterns (Figure 4). Correspondingly, we conducted a latent profile analysis to understand how usage patterns clustered.

**Figure 4.**
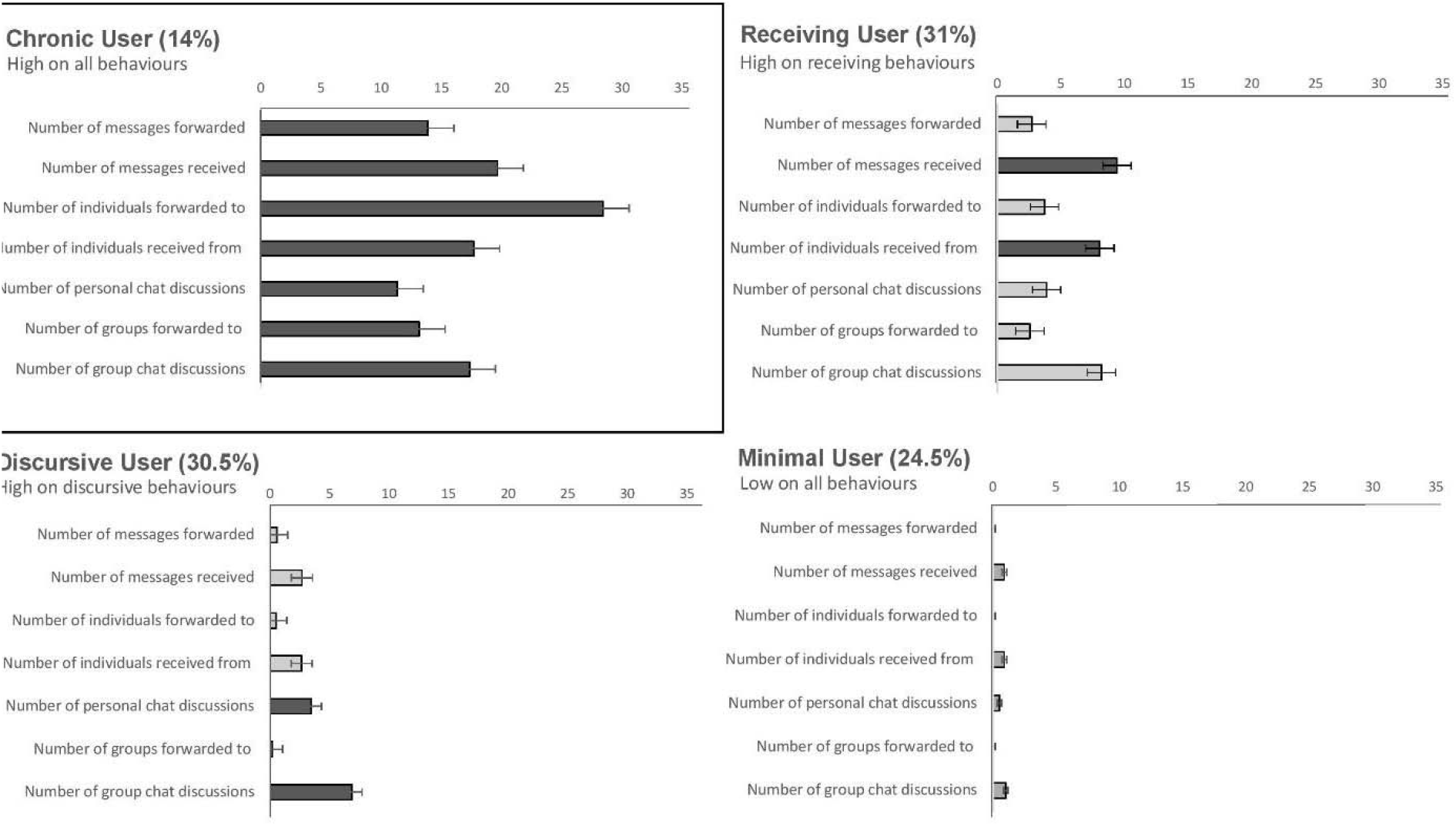
Taxonomy of COVID-19 related WhatsApp usage. Using latent profile analysis, participants were classified based on how they had used WhatsApp for COVID-19 content during a week of monitoring. The figure depicts WhatsApp usage activities for the chronic user (top left), receiving user (top right), discursive user (bottom left), and minimal user (bottom right). Horizontal lines represent the 95% confidence interval for the mean.

A 4-cluster solution yielded the lowest absolute BIC values (Multimedia Appendix 2), resulting in the following taxonomy (Figure 4). First, 14% (21/151) of participants were “chronic users” with high activity in each of the WhatsApp usage variables. Correspondingly, this group of participants was responsible for receiving and transmitting a large volume of forwarded COVID-19 messages, sending the messages both to individual contacts and to groups. Second, 31% (47/151) of participants were “receiving users” distinguished by their receipt of multiple COVID-19 forwarded messages. Although this group discussed COVID-19 frequently in group chats, they rarely passed along forwarded COVID-19 messages they had received. A third group, “discursive users” (31%, 46/151), had low exposure to forwarded COVID-19 messages and primarily engaged with COVID-19 content through personal and group chats. Finally, 25% (37/151) of participants were “minimal users” with low engagement with COVID-19 content overall.

#### Understanding user characteristics

As an exploratory analysis, we performed a classification tree analysis to predict WhatsApp user type based on: demographics, COVID-19 concerns, depression and anxiety scores (DASS-21), and the time of day for WhatsApp usage. We applied recursive partitioning (“rpart”), a machine learning technique that allows multiple variables to be analyzed simultaneously, and supports the modelling of complex, non-linear relations between predictors [23]. To avoid overfitting, the final tree was pruned by selecting the tree size with the lowest cross-validation error (minimized with a tree size of 8 for our dataset).

As shown in Figure 5, chronic users were more likely to be married / divorced, and to send messages either throughout the day or in the afternoon. In terms of COVID-19 responses, chronic users either had: (i) extreme fears of the COVID-19 situation (low or high), or (ii) had moderate fears paired with lower confidence in the government’s response (low or moderate).

**Figure 5.**
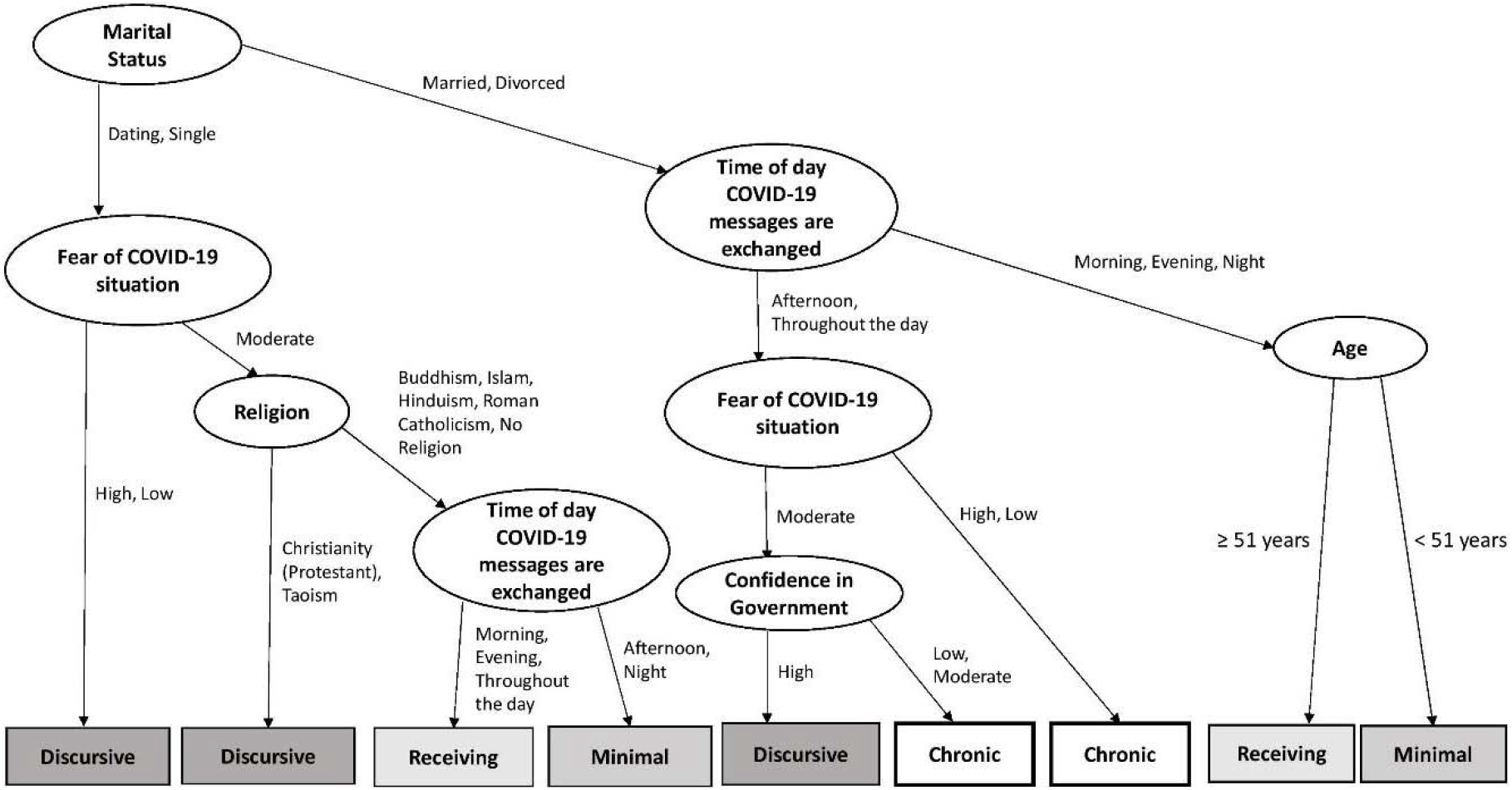
Classification tree analysis. Recursive partitioning was used to predict which of 4 WhatsApp usage profiles (chronic, receiving, discursive, or minimal) participants belonged to, based on baseline questionnaire measures (demographics, COVID-19 concerns, scores on the Depression, Anxiety and Stress Scale, and time of WhatsApp usage). The final tree model is presented as a flow chart, with factors chosen at each level to categorize the maximal number of participants. Marital status, time of WhatsApp usage, and age emerged as the primary predictors (model classification accuracy: 64.2%, above the chance level of 25%).

On the other hand, discursive users were more likely to be single / dating, and had either: (i) extreme levels of COVID-19 fears (either high or low), or (ii) moderate fear levels alongside Christian or Taoist affiliations. A sub-group of discursive users were – like chronic users – married / divorced, with moderate levels of COVID-19 fears. However, they were distinguished from chronic users by their high confidence in government (as compared to chronic users’ lower confidence).

Finally, receiving and minimal users had similar profiles. If single / dating, both sets of users tended to have moderate levels of COVID-19 fears, had a wide range of religious backgrounds, and were distinguished by the time of day that COVID-19 related messages were received (receiving users: morning, evening, and throughout the day; minimal users: afternoon and night). If married / divorced, both sets of users tended to send messages at only one time of the day (morning, evening, or night), and were distinguished by age (receiving users: 51 years and above; minimal users: below 51 years). Table 1 describes the demographic characteristics of the four profiles.

#### Does WhatsApp usage relate to COVID-19 concerns?

As the final thrust of the study, we ran linear mixed effects models to examine whether WhatsApp usage related to COVID-19 concerns (Table 2). As shown in Figure 6, day-to-day COVID-19 fears and thoughts fluctuated (*t*(249.13) = -3.72, *P* < 0.001 ; *t*(297.02) = -2.36, *P* = 0.02).

**Table 2.** Parameter estimates for the multi-level model of thoughts about COVID-19 (Model 1) and fear of COVID-19 (Model 2) as a function of participants’ daily WhatsApp use (personal-chat and group-chat).

| [Model 1] Outcome: Thoughts about COVID-19 |  |  |  |  |  |  |
| --- | --- | --- | --- | --- | --- | --- |
| Fixed effects | Estimate | SE | *t* | *P* | CI<sub>95</sub><br>lower | CI<sub>95</sub><br>upper |
| Intercept | 2.18 | 0.07 | 32.81 | <0.001 | 2.05 | 2.31 |
| Time (centered) | -0.03 | 0.01 | -2.36 | 0.02 | -0.05 | -0.00 |
| Daily personal chat usage (between) | 0.04 | 0.02 | 2.36 | 0.02 | 0.00 | 0.07 |
| Daily personal chat usage (within) | 0.00 | 0.01 | 0.42 | 0.68 | -0.01 | 0.02 |
| Daily group chat usage (between) | 0.05 | 0.06 | 0.89 | 0.37 | -0.06 | 0.17 |
| Daily group chat usage (within) | -0.00 | 0.03 | -0.08 | 0.93 | -0.06 | 0.05 |
| Random effects | Estimate | SE | *Z* | *P* | CI<sub>95</sub><br>lower | CI<sub>95</sub><br>upper |
| Intercept (between) | 0.56 | 0.08 | 6.89 | <0.001 | 0.42 | 0.75 |
| Residual (within) | 0.37 | 0.02 | 14.90 | <0.001 | 0.33 | 0.43 |
| Autocorrelation (within) | 0.24 | 0.05 | 4.97 | <0.001 | 0.14 | 0.33 |
| [Model 2] Outcome: Fear of COVID-19 |  |  |  |  |  |  |
| Fixed effects | Estimate | SE | *t* | *P* | CI<sub>95</sub><br>lower | CI<sub>95</sub><br>upper |
| Intercept | 2.10 | 0.06 | 36.37 | <0.001 | 1.98 | 2.21 |
| Time (centered) | -0.03 | 0.01 | -3.72 | <0.001 | -0.05 | -0.02 |
| Daily personal chat usage (between) | 0.01 | 0.01 | 0.85 | 0.39 | -0.02 | 0.04 |
| Daily personal chat usage (within) | 0.01 | 0.01 | 1.22 | 0.24 | -0.01 | 0.02 |
| Daily group chat usage (between) | 0.02 | 0.05 | 0.49 | 0.62 | -0.07 | 0.12 |
| Daily group chat usage (within) | -0.03 | 0.02 | -1.42 | 0.17 | -0.06 | 0.01 |
| Random effects | Estimate | SE | *Z* | *P* | CI<sub>95</sub><br>lower | CI<sub>95</sub><br>upper |
| Intercept (between) | 0.44 | 0.06 | 7.47 | <0.001 | 0.34 | 0.58 |
| Residual (within) | 0.21 | 0.01 | 13.83 | <0.001 | 0.18 | 0.23 |
| Autocorrelation (within) | 0.26 | 0.05 | 5.16 | <0.001 | 0.16 | 0.35 |

**Figure 6.**
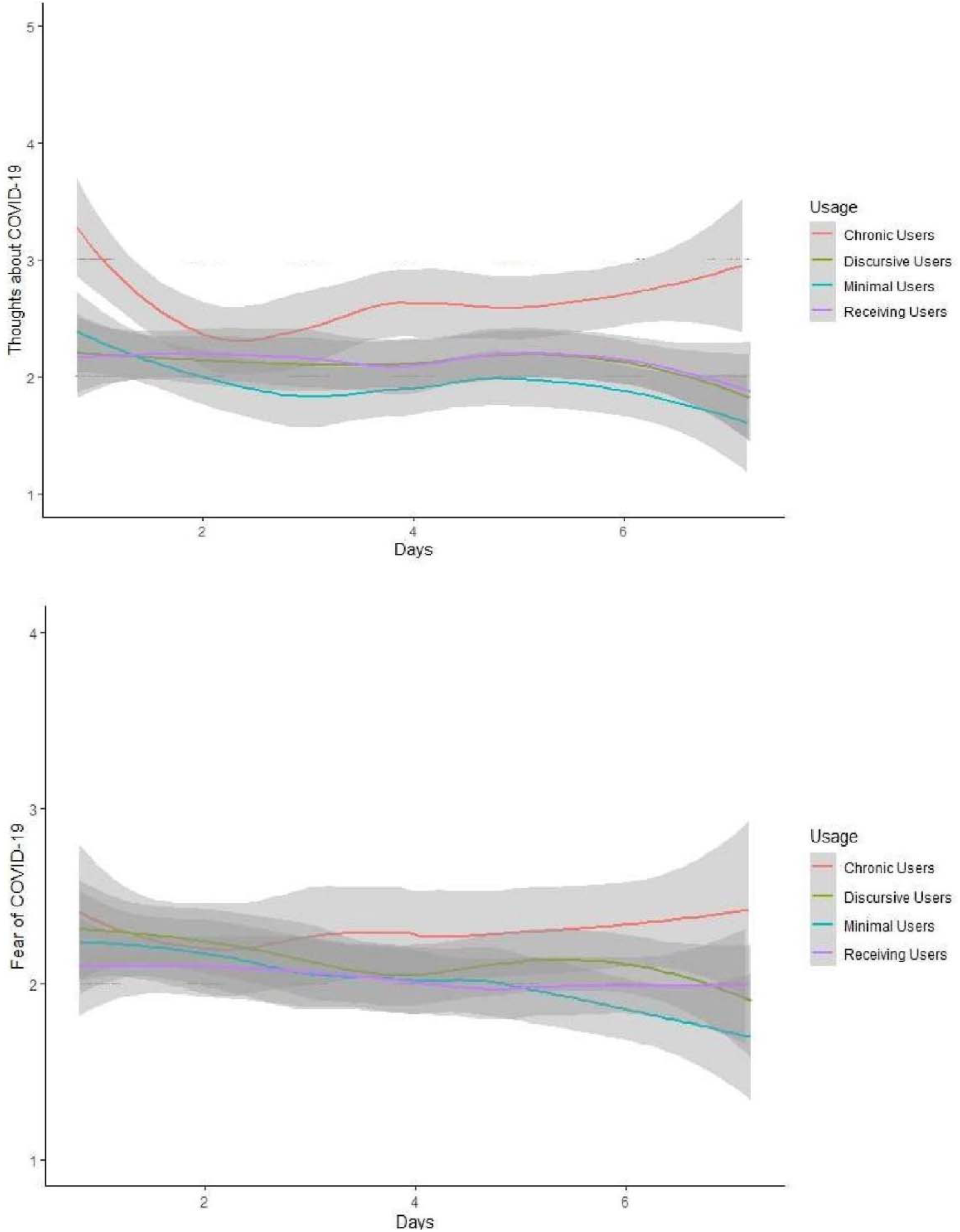
COVID-19 related thoughts and fears over a week. Day-to-day variations in COVID-19 thought (top) and fear levels (bottom), as a function of WhatsApp user profiles. The shaded grey areas represent 95% confidence intervals.

For COVID-19 thoughts, there was a significant effect of WhatsApp personal chat usage at a between-subjects level; *t*(164.81) = 2.36, *P* = .02). That is, participants who handled higher levels of COVID-19 content in their personal chats reported having more COVID-19 thoughts (relative to participants who handled lower levels of COVID-19 content). However, the corresponding effect for group chats was not significant; *t*(141.17) = 0.89, *P* = .37. At the within-subjects level, neither day-to-day fluctuations in personal nor group chat activities significantly predicted COVID-19 thoughts (smallest *P* = .68).

For COVID-19 fears, we found no significant effect of any WhatsApp usage variable (smallest *P* = .17). (For sensitivity analyses, we repeated both models with group membership as a fixed factor in place of personal and group chat usage, and our primary conclusions did not change; Multimedia Appendix 3.)

## Discussion

The ongoing pandemic has drawn attention to the role of social media in public health. Against this backdrop, we present the first infodemiological study documenting the spread of COVID-19 content through WhatsApp. By tracking daily WhatsApp usage for one week, we found that: (i) nearly every participant engaged in “COVID-19 chatter”, and that (ii) participants were more likely to share or receive forwarded messages than to engage in conversation about COVID-19.

Our findings have several implications for health communication. First, the volume of forwarded messages we observed raises concern. On other social media platforms, forwarding behaviors have been linked to the spread of misinformation. For example, a study of 4.5 million Twitter posts found that misinformation was 70% more likely to be shared than posts that were true; correspondingly, any single re-tweet had a higher probability of containing false than truthful news [15]. Although analogous research has not been conducted on WhatsApp, the app’s developers have likewise targeted forwarded messages as being at high-risk for misinformation [4,5].

To the extent that forwarded messages carry misinformation [1, 2], our latent profile analyses revealed that 1 in 10 participants were ‘chronic users’ who received and shared a large volume of these messages. Notably, chronic users disseminated an average of 14 forwarded messages during the week – approximately 5 times the number of messages sent by all participants in the study. This is reminiscent of research from other social media platforms (e.g., Twitter), where a small group of “supersharers” and “superconsumers” are responsible for the bulk of misinformation shared [26]. Given the potential influence of this group, further research is needed to understand: (i) the profile of chronic users, (ii) the reasons why they forward messages, and (iii) how their forwarding activities may influence outcomes during health crises.

Aside from chronic users, our study also found that 1 in 3 participants were ‘receiving users’ who had high exposure to forwarded COVID-19 content. Receiving users tended to be older (in line with misinformation studies on Facebook [27]) but were otherwise moderate in profile - whether in terms of COVID-19 fears or in religion (coming from a diverse religious background). Although this group did not spread forwarded messages themselves, their high exposure may nonetheless leave them susceptible to false beliefs. Again, we urge further research to understand how WhatsApp use within this group influences their health behaviors during crises.

Taken together, our taxonomy of WhatsApp user profiles provides a basis for targeted risk communication. Our findings suggest that public health agencies may need to reach out proactively to chronic and receiving users who handle the bulk of forwarded COVID-19 content on WhatsApp. One possible intervention may be to encourage these users to subscribe to official WhatsApp channels for updates (e.g., from the World Health Organization) [2], capitalizing on their pre-existing readiness to use the platform.

Finally, aside from forwarded content, we also found that WhatsApp users who discussed COVID-19 in their personal chats were more likely to think about COVID-19 through the day. As similar forms of rumination (involving frequent and persistent thoughts) have been linked to clinical depression [24], this finding may implicate COVID-19 chatter as a risk factor for poorer well-being [25]. Future studies should explore this possibility and the potential mechanisms involved.

### Limitations

In reporting these findings, we note several limitations. First, we opted to study WhatsApp, the most widely used messenger application. At this juncture, it is unclear whether our results will generalize to other messenger applications (e.g., Facebook Messenger, Telegram).

Second, our recruitment strategy was limited by the nature of the pandemic. Owing to infectious disease protocols and the short time period when crisis-related communication was high [28], we relied on online recruitment. Further research is needed to examine whether our findings generalize to the broader population.

Finally, although the experience sampling method captured WhatsApp usage in participants’ naturalistic settings, the method nonetheless required self-reports. Extending our findings, future studies will profit from having objective metrics of WhatsApp usage.

## Conclusions

In conclusion, we applied the experience sampling method to capture for the first time “COVID-19 chatter” on WhatsApp. In total, we tracked 924 days of chatter *in situ*, revealing: (i) the sheer prevalence of WhatsApp usage; (ii) a typology of WhatsApp users; and (iii) a link between usage patterns and constant thoughts about the pandemic. These findings bring the field one step closer to digital phenotyping through WhatsApp, spurring future research on the role of messenger applications in health communication.

## Data Availability

The data that support the findings of this study are available on request from the corresponding author (JCJL). The data are not publicly available due to IRB restrictions.

## Acknowledgements

RREW, KJQH, JWEC contributed to the study design and collected the data. EYQT and SYE analyzed the data and wrote the first draft of the manuscript. JCJL and EMWT conceived of the study, had oversight of the overall project, and supervised both data analyses and manuscript preparation. All authors provided feedback on the final version of the manuscript. This research was funded by a grant awarded to JCJL from the NUS Centre for Trusted Internet and Community (Grant number: CTIC-RP-20-09). The first author’s (EYQT) involvement was funded by a centre grant awarded to the Centre for Sleep and Cognition. This study was approved by the Yale-NUS College Ethics Review Committee.

## Abbreviations

COVID-19: Coronavirus disease
GMM: Gaussian Mixed Models
BIC: Bayesian Information Criterion
ICL: Integrated Completed Likelihood Criterion
BLRT: Bootstrap Likelihood Test

