## Supplementary figures and images for "Tracking private WhatsApp discourse about COVID-19: A longitudinal infodemiology study in Singapore"

### Supplemental Figure 1

#### Appendix A

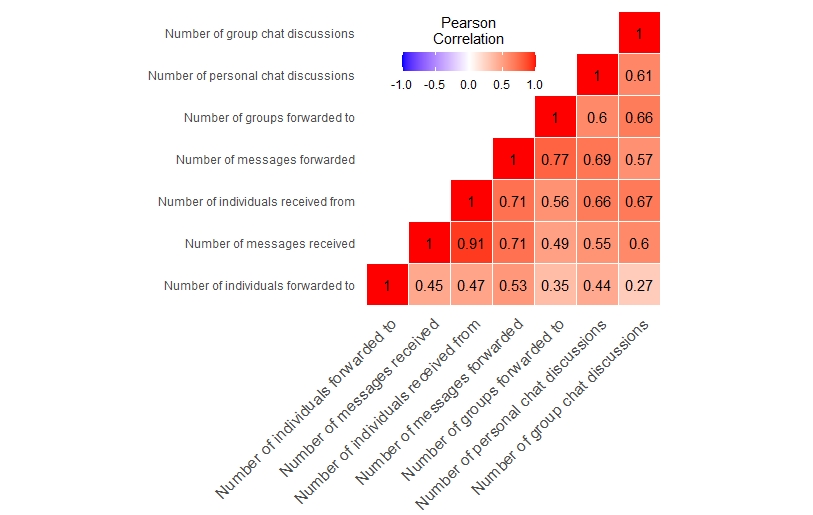

**Figure S1.** Correlation matrix of the 7 quantitative WhatsApp usage variables.
