## Supplemental Table 2 for "Tracking private WhatsApp discourse about COVID-19: A longitudinal infodemiology study in Singapore"

### Appendix B

**Table S2. Model fit indices.** Model fit indices for Latent Profile Analysis. Lower

absolute values indicate better model fit.

| Solution | BIC | ICL | BLRT (*p*) |
| --- | --- | --- | --- |
| 1 cluster | -2313.26 | -2313.26 |  |
| 2 clusters | -1081.72 | -1081.82 | 1382.06 (0.001) |
| 3 clusters | -614.58 | -616.65 | 617.65 (0.001) |
| 4 clusters | -466.96 | -468.20 | 298.14 (0.001) |
| 5 clusters | -575.41 | -579.63 | 42.07 (0.78) |

For sensitivity analyses, we used ICL and obtained similar results. Likewise, BLRT indicated that the 4-cluster solution was a better fit than a 1-cluster solution (P = .001), 2-cluster solution (P = .001), and 3-cluster solution (P = .001). As there was no statistical difference between a 5- and 4-cluster solution (P = .78), the 4-cluster solution provided the optimal fit based on parsimony (see Figure S2).

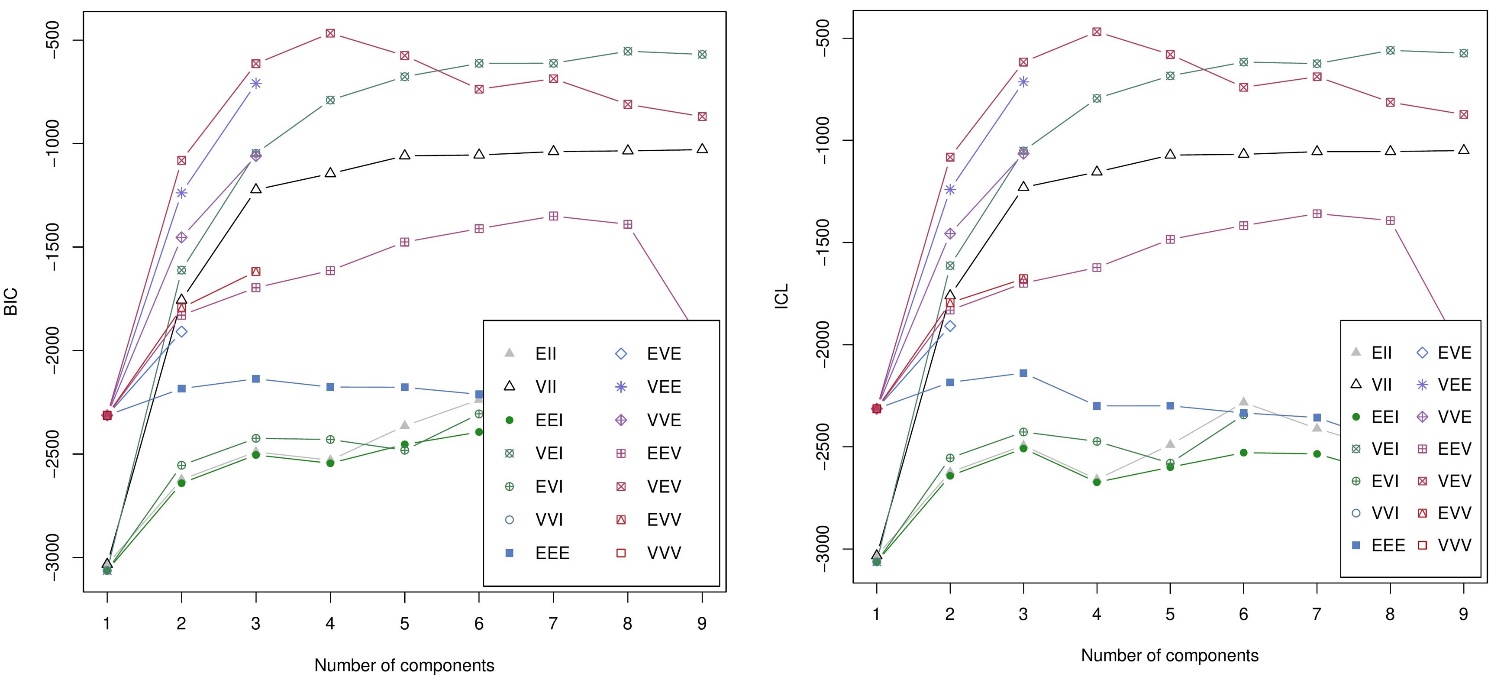

**Figure S2.** **BIC and ICL plots.** The optimal number of clusters was determined using the **(A)** Bayesian Information Criterion (left) and **(B)** Integrated Completed Likelihood Criterion (right). Lower absolute BIC and ICL values represented better model fit.
