## Supplemental Table 3 for "Tracking private WhatsApp discourse about COVID-19: A longitudinal infodemiology study in Singapore"

### Appendix C

**Table S3. Model parameter estimates (group membership).** Parameter estimates for the multi-level model of thoughts about COVID-19 (Model 1) and fear of COVID-19 (Model 2) as a function of participants’ group membership.

| [Model 1] Outcome: Thoughts about COVID-19 | |  |  |  |  |  | |
| --- | --- | --- | --- | --- | --- | --- | --- |
| Fixed effects | Estimate | SE | t | P | | CI_95_  lower | CI_95_  upper |
| Intercept | 2.19 | 0.15 | 14.41 | <0.001 | | 1.89 | 2.49 |
| Time (centered) | -0.03 | 0.01 | -2.51 | 0.01 | | -0.06 | -0.01 |
| Group membership | 0.00 | 0.06 | 0.01 | 0.99 | | -0.12 | 0.12 |
| Random effects | Estimate | SE | Z | P | | CI_95_  lower | CI_95_  upper |
| Intercept (between) | 0.57 | 0.09 | 6.64 | <0.001 | | 0.42 | 0.76 |
| Residual (within) | 0.42 | 0.03 | 13.42 | <0.001 | | 0.37 | 0.49 |
| Autocorrelation (within) | 0.32 | 0.05 | 6.23 | <0.001 | | 0.22 | 0.42 |
| [Model 2] Outcome: Fear of COVID-19 |  |  |  |  | |  |  |
| Fixed effects | Estimate | SE | t | P | | CI_95_  lower | CI_95_  upper |
| Intercept | 2.05 | 0.13 | 15.88 | <0.001 | | 1.80 | 2.31 |
| Time (centered) | -0.04 | 0.01 | -3.93 | <0.001 | | -0.05 | -0.02 |
| Group membership | 0.020 | 0.05 | 0.43 | 0.67 | | -0.08 | 0.12 |
| Random effects | Estimate | SE | Z | P | | CI_95_  lower | CI_95_  upper |
| Intercept (between) | 0.44 | 0.06 | 7.47 | <0.001 | | 0.34 | 0.58 |
| Residual (within) | 0.22 | 0.01 | 15.23 | <0.001 | | 0.19 | 0.25 |
| Autocorrelation (within) | 0.25 | 0.49 | 5.14 | <0.001 | | 0.15 | 0.34 |
